# Increased risk of death in covid-19 hospital admissions during the second wave as compared to the first epidemic wave. A prospective dynamic cohort study in South London, UK

**DOI:** 10.1101/2021.06.09.21258537

**Authors:** Martina Cusinato, Jessica Gates, Danyal Jajbhay, Tim Planche, Yee-Ean Ong

## Abstract

**Objective:** To assess whether mortality of patients admitted for covid-19 treatment was different in the second UK epidemic wave of covid-19 compared to the first wave accounting for improvements in the standard of care available and differences in the distribution of risk factors between the two waves.

**Design:** Single-centre, analytical, dynamic cohort study.

**Participants:** 2,701 adults (≥18 years) with SARS-CoV-2 infection confirmed by polymerase chain reaction (PCR) and/or clinico-radiological diagnosis of covid-19, who required hospital admission to covid-19 specific wards, between January 2020 and March 2021. There were 884 covid-19 admissions during the first wave (before 30 Jun 2020) and 1,817 during the second wave.

**Outcome measures:** in-hospital covid-19 associated mortality, ascertained from clinical records and Medical Certificate Cause of Death.

**Results:** The crude mortality rate was 25% lower during the second wave (2.23 and 1.66 deaths per 100 person-days in first and second wave respectively). However, after accounting for age, sex, dexamethasone, oxygen requirements, symptoms at admission and Charlson Comorbidity Index, mortality hazard ratio associated with covid-19 hospital admissions was 1.62 (95% confidence interval 1.26, 2.08) times higher in the second wave compared to the first.

**Conclusions:** Analysis of covid-19 admissions recorded in St. Georges Hospital, shows a larger second epidemic wave, with a lower crude mortality in hospital admissions. Nevertheless, after accounting for other factors underlying risk of death for covid-19 admissions was higher in the second wave. These findings are temporally and ecologically correlated with an increased circulation of SARS-CoV-2 variant of concern 202012/1 (alpha).

## INTRODUCTION

Since its emergence in December 2019, the spread of SARS-CoV-2 has increased exponentially leading to the declaration of a pandemic by the World Health Organization (WHO) on 11 March 2020, marking the beginning of an outbreak that has posed immense challenges for health care systems across the globe [1]. The first confirmed case in the United Kingdom (UK) was registered on 31 January 2020. At the beginning of March 2020, the growing transmission rates lead to the introduction of a series of control measures that escalated to a full national lockdown (23 March 2020). This was subsequently followed by a drop in transmission and hospitalisation rates with restrictions being eased over the summer months (Jun – Aug 2020). However, in October 2020 infections began to increase again leading to a second wave of covid-19 cases. The implementation of a second lockdown (04 November 2020) followed by tiered control measures, in place until the beginning of March 2021, were needed to reduce the transmission rates again [2]. As of 01 May 2021, the UK has recorded 4,418,819 confirmed cases, 463,485 hospital admissions, and 127,571 deaths [3].

During the first wave of covid-19, relatively little was known about this novel illness and management was largely based upon experience of treating other viral infections. However, since the start of the pandemic a great deal has been learned about treatment of covid -19 and there have been several important changes to the management of patients admitted with covid -19. From the start of the second wave, dexamethasone was prescribed to all patients requiring supplemental oxygen and Remdesivir was administered to hypoxemic patients presenting within 10 days of symptoms onset. The indications for Tocilizumab changed during the second wave where patients initially had access to this only within clinical trials. Differences in the management and standard of care across waves need to be accounted for when analysing covid-19 mortality over time [4-8].

A key feature of the second wave of covid-19 in the UK, was the emergence of a new SARS-CoV-2 variant designated VOC 202012/01 or alpha (lineage B.1.1.7). This new variant was identified in December 2020 by Public Health England through genomic sequencing of samples originally taken in south east England in early October 2020. Since then, VOC 202012/01 has become the predominant variant circulating in the UK [9-11]. Several studies have established that VOC 202012/01 is more transmissible than pre-existing variants but its impact on mortality has not been widely studied. To our knowledge, three studies have been recently published, reporting an increase in mortality among those with a positive test in the community; however, the impact on in-hospital mortality remains poorly understood [12-14].

St George’s University Hospital NHS Foundation Trust is one of the largest hospitals in the UK and is based in South West London. It serves a local catchment population of 560,000 and specialist services to 3.4 million people. The objective of this study is to assess whether mortality of patients admitted for covid-19 treatment was different in the second UK wave of covid-19 compared to the first wave accounting for differences in the standard of care available in each wave.

## METHODS

### Study design

This is a single-centre, analytical, dynamic cohort study using data extracted from routinely collected, electronic medical records and hospital database.

### Participants and setting

The study population for this cohort study comprised all adults (≥18 years) with SARS-CoV-2 infection confirmed by polymerase chain reaction (PCR) and/or clinico-radiological diagnosis of covid-19, who required hospital admission to covid-19 specific wards at St George’s University Hospitals NHS Foundation Trust (London, UK). Patients seen in the Emergency Department or in Acute Medical Units (AMU) who were discharged on the same day were not included. Although covid-19 wards opened in March 2020, the study period encompasses admissions between 01 Jan 2020 and 31 March 2021, as some of the early patients admitted to covid-19 wards were already hospitalised. All patients meeting the inclusion criteria during the study period were included in the cohort. There was no a priori study size calculation.

### Data sources and measurement

The study cohort was identified retrospectively using hospital records of admissions to active covid-19 wards. These lists included patient identifiers, hospital admission date, ward and administrative information. Respiratory and Intensive Care clinicians within the study team and involved in the care of covid-19 patients, reviewed the electronic medical records for all the patients in the initial list, confirming criteria for covid-19 admission. In case of multiple covid-19 admissions, only the most severe, as defined by the highest respiratory support needed, was included [15].

Study follow-up (from admission to discharge/outcome) was also carried out by clinicians, prospectively, through review of electronic medical records. Patient data was extracted manually using a standardised electronic questionnaire and was supervised by a senior clinician within the Respiratory team. These data were also obtained through the informatic department and linked using hospital identifiers (laboratory, pathology results and ethnicity data).

The follow-up period for this study began at admission and ended at outcome occurrence (death) or censoring. Participants were censored at hospital discharge or at 6 months if admissions exceeded this period (1 patient only).

PCR pathology results were available for all tests requested during the study period, so we matched these with our cohort of patients. Those with positive PCR results dated at least 15 days after their hospital admission were considered probable hospital acquired infections (HAI) and had the start of their follow-up (time at risk) amended to be 14 days (maximum incubation period [16]) before the date of the positive PCR result, instead of the actual admission day.

### Variables

The outcome variable was in-hospital covid-19-associated mortality, ascertained from clinical records and Medical Certificate Cause of Death (MCCD). The main explanatory variable for this analysis was covid-19 wave, and 31 June 2020 used as cut-off to separate both waves. First wave was used as baseline/reference. Covariates of interest for this analysis included demographics (sex, age at admission, ethnicity), symptoms at admission, Body Mass Index (BMI), treatment (dexamethasone, Remdesivir, Tocilizumab), oxygen requirement, HFNO/CPAP (High Flow Nasal Oxygen/Continuous Positive Airway Pressure), invasive ventilation, Intensive Care Unit (ICU) admission, Clinical Frailty Score (CFS), Charlson Comorbidity Index (CCI), social history. Most variables were used in their original scale, others were recategorised using clinically relevant categories with a sufficient number of participants in each group to avoid sparsity.

Where categorised, age groups in years were: [18,40), [40,60), [60,80), ≥80. BMI at admission was grouped using categories derived from the WHO classification of BMI (in kg/m^2^): <18.5, [18.5,25), [25,30), ≥30[17]. Oxygen requirement was a dichotomous variable indicating whether the maximum FiO2 (Fraction of Inspired Oxygen) was over 21%. ICU admission was defined as covid-19 pneumonitis admitted to ICU. Symptoms at admission were respiratory or wider infective symptoms at time of presentation. The CFS level was collected on a nine-point ordinal scale to assess frailty within two weeks of admission (1 being very fit, 2 well, 3 managing well, 4 vulnerable, 5 mildly frail, 6 moderately frail, 7 severely frail, 8 very severely frail, and 9 terminally ill); but to avoid sparsity categories 7 to 9 were grouped [18]. CFS was expanded to include all age groups excepting those patients with disabilities which rendered it inappropriate. CCI is a widely used comorbidity summary measure, based on age and a predefined number of conditions with an assigned integer weight representing the severity of each condition; for this analysis scores of 8 or more were grouped in one category [19]. All scores were calculated by clinicians experienced in the use of scales. Four categories under social history reflected the level of assistance required for daily activities.

### Statistical methods

The distribution of covariates was assessed for the entire cohort and across waves. Mortality rates and person-time of observation were calculated for the main exposure groups and all covariates of interest. The strength of the association was quantified using incidence rate ratios (IRR), and the statistical significance using 95%CIs and p-values. Survival across the different waves was explored using time-to-event analysis and log-rank to test the significance of the difference between the survival curves.

Cox regression was used to estimate the effect of wave on mortality adjusting for multiple covariates. The proportional hazard assumption was explored graphically and by testing for a zero slope in Schoenfeld residuals. Follow-up time was stratified using lexis expansion and to minimise bias, intervals were created so they would contain the same number of events. The assumption of proportionality was supported.

A causal model was built using a stepwise backward approach where (non-forced) pre-defined covariates were retained in the model unless there were problems with multicollinearity. Age and gender were considered *a priori* confounders (forced variables). Age was fitted using restricted cubic splines, with knots positioned so numbers of events between knots were approximately equally distributed. The full model included age, gender and all variables found to confound the crude association between wave and mortality (non-forced variables). A change in the magnitude ≥ 5.5% was considered an indication of confounding. ICU admission was included in the model as a non-forced variable regardless of the degree of confounding of the main association. Problems will multicollinearity on the main effect in the full model, were resolved using RMSE (Root Mean Square Error) reduction for backward deletion of non-forced variables. The RMSE for the full model was used as reference for each step; and RMSE for each reduced model was calculated as √[*⟦ ⟦* (*β ⟧*_(1 *reduced*) – *β*_(1 *full*))*⟧* ^2+ *⟦SE⟧*_*reduced*^2] [20].

Following the same methodology, we carried out a sub-analysis among those requiring ICU admission. Data management and statistical analysis were carried out using R.

### Governance and ethics

This study was approved by the Health Research Authority (20/SC/0220). This manuscript follows the STROBE statement for reporting of cohort studies.

## RESULTS

Between 01 January 2020 and 31 March 2021, there were 3,376 covid-19 positive adult patients registered at St. Georges Hospital. Of these, 2,701 were patients admitted to covid-19 wards for treatment, all of whom were included. 32.7% (884) in the first wave and 67.3% (1,817) in the second wave (Figure 1). At the time of database cut-off (23 May 2021), there were 16 patients with no outcome recorded.

**Figure 1:**
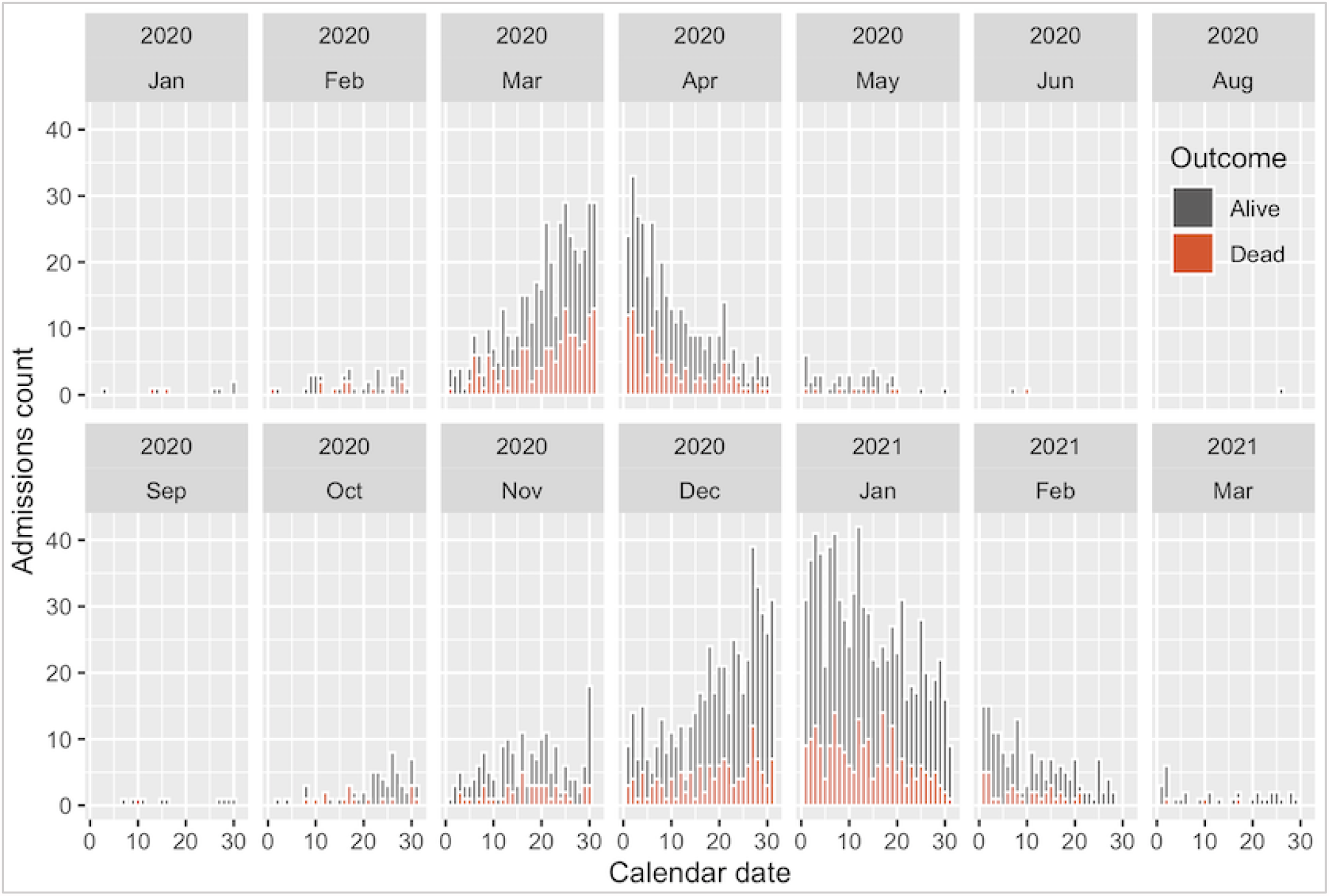
Number of admissions per day according to outcome.

The distribution of characteristics at admission for the entire cohort and across waves is shown in Table 1. Covid-19 patients admitted during the second wave, were more likely to be younger, with patients aged 40 to 60 years being more prevalent in the second wave (495, 27.2%) and patients aged over 80 years being more prevalent in the first wave (273, 30.9%). The distribution of sex was similar for both waves with males being overrepresented (57.6%). During the second wave, patients were more likely to lead an independent life (1,101, 61.1%) or have some level of family assistance (369, 20.5%); intermediate levels of frailty (3 managing well, 4 vulnerable, 5 mildly frail) were also more prevalent in the second wave (1,103, 60.7%) than in the first wave (392, 44.3%). The proportion of severely frail patients (CFS 7-9) admitted was lower in the second wave (197, 10.8% vs. 164, 18.6% in the first wave). Admissions scoring 0-3 in CCI were more prevalent during the second wave (891, 49.0% vs. 386, 43.7%); the reverse occurred for CCI scores 4-5 and over 6 (29.3% and 27.0% respectively for first wave, vs. 25.3% and 25.7% for second wave). Absence of respiratory or wider infective symptoms at onset (initial diagnosis through PCR) was more prevalent in the second wave (361, 19.9%) compared to the first wave (50, 5.7%).

**Table 1:**
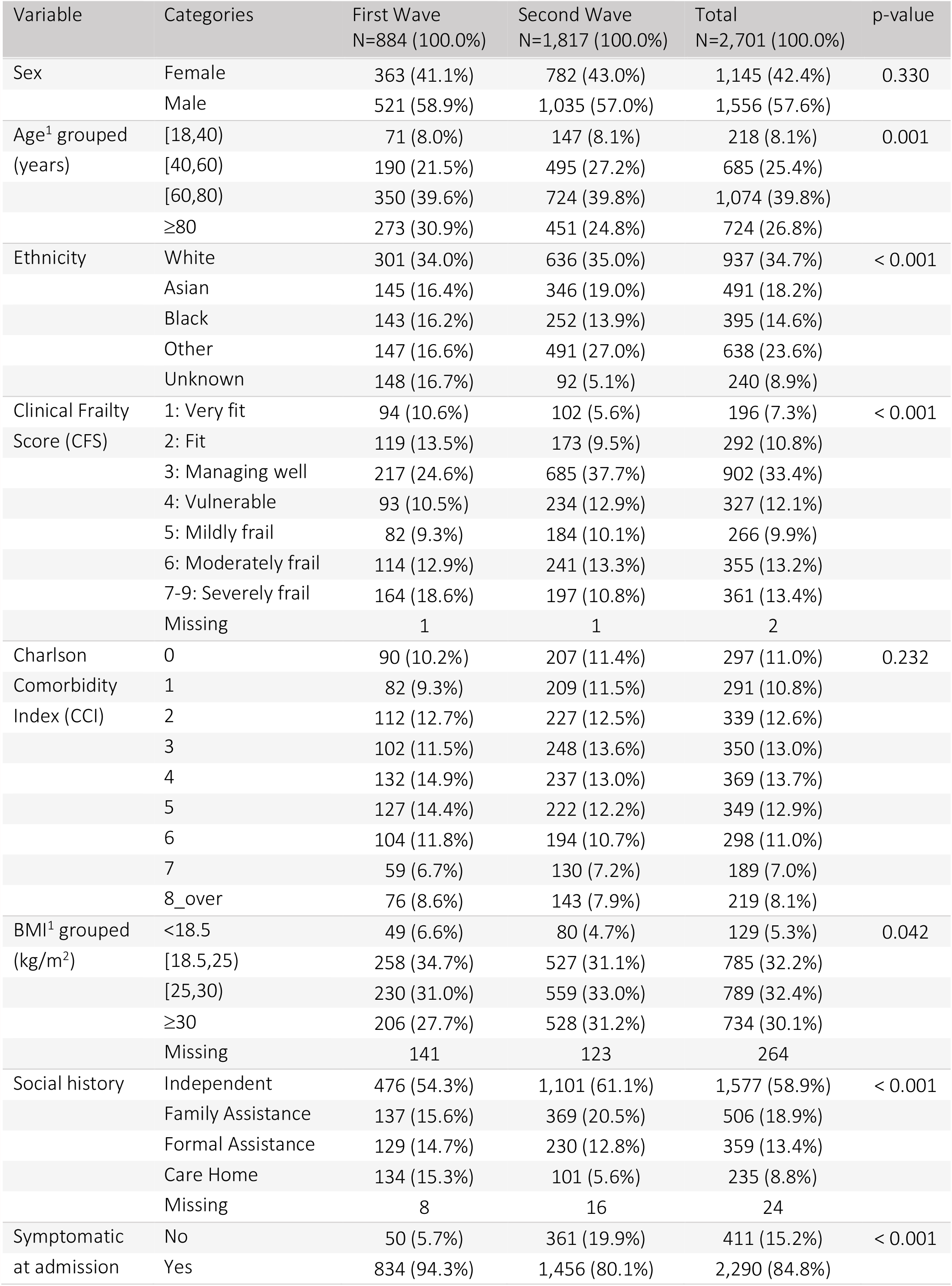
Baseline characteristic of the study population and comparison groups

The distribution of medical interventions after admission are listed in Table 2. The prevalence of admitted patients requiring oxygen during admission was similar in both waves: 76.3% (668) in the first wave and 73.5% (1,328) in the second wave. The use of HFNO/CPAP was more prevalent in the second wave (400, 22.2%) than during the first wave (81, 9.3%), whilst invasive ventilation was more prevalent in the first wave. The distribution of patients requiring ICU admission had a similar distribution across waves (23.1%). The use of dexamethasone, Remdesivir and Tocilizumab was almost exclusively during the second wave. During the first wave there was some use of Tocilizumab (23, 2.6%) and dexamethasone (58, 6.6%), both used in the context of clinical trials (with a few cases additional cases of compassionate use for Tocilizumab).

**Table 2:** Distribution of medical interventions after admission across waves

| Variable / Categories | First Wave<br>N=884 (100.0%) | Second Wave<br>N=1,817 (100.0%) | Total<br>N=2,701 (100.0%) | p-value |
| --- | --- | --- | --- | --- |
| ICU admission | 211 (23.9%) | 412 (22.7%) | 623 (23.1%) | 0.489 |
| Oxygen requirement | 668 (76.3%) | 1,328 (73.5%) | 1,996 (74.4%) | 0.118 |
| Oxygen requirement missing | 9 | 11 | 20 |  |
| HFNO/CPAP | 81 (9.3%) | 400 (22.2%) | 481 (18.0%) | < 0.001 |
| HFNO/CPAP Missing | 9 | 18 | 27 |  |
| Invasive ventilation | 178 (20.3%) | 237 (13.1%) | 415 (15.5%) | < 0.001 |
| Invasive ventilation Missing | 6 | 10 | 16 |  |
| Dexamethasone | 58 (6.6%) | 1,241 (68.3%) | 1,299 (48.1%) | < 0.001 |
| Tocilizumab | 23 (2.6%) | 229 (12.6%) | 252 (9.3%) | < 0.001 |
| Remdesivir | 0 (0.0%) | 575 (31.6%) | 575 (21.3%) | < 0.001 |

(1) Brackets indicate that the side of the interval is closed; parenthesis, that the number is excluded.
ICU: Intensive Care Unit, HFNO/CPAP: High Flow Nasal Oxygen/Continuous Positive Airway Pressure

A total of 752 patients died over the total time at risk (40,777 person-days); 297 (33.6%) deaths occurred during the first wave and 455 (25.3%) during the second wave. The median time of follow-up for those discharged was 10 days (IQR: 5-22 days) and for those who died was 11 days (IQR: 5-19 days). We found no differences in the overall distribution of the follow-up time across waves. Among those discharged, admissions with lengths of stay (LOS) over 35 days were similar across waves (11.9% for the first wave and 11.1% for the second); LOS between 0-7 days were more prevalent in the second wave (562, 41.8%) than in the first wave (188, 32.0%). The median probability of survival was 29 days (95%CI 30-41 days) for the first wave, and 37 days (95%CI 32-47 days) for the second.

In this cohort, patients admitted during the second wave of the covid-19 pandemic, had a (crude) mortality rate 25% lower than that of patients admitted during the first wave (IRR 0.75, 95%CI 0.64, 0.86). Mortality rates, and crude IRR for all variables of interest are shown in table 1 of the supplemental materials. Overall, mortality rates were 1.19 times higher in men than in women (95%CI 1.03, 1.38) and 1.37 times higher in patients of Asian ethnicity compared to white ethnicity (95%CI 1.12, 1.67). Crude IRR was 1.92 times higher (95%CI 1.27, 3.09) among those aged 60-79 years, and 2.81 times (95%CI 1.85, 4.50) in those aged over 80 years compared to patients younger than 40 years. Mortality rates for patients with some degree of family assistance was 1.69 higher (95%CI 1.42, 2.02) than those living an independent life. Further, IRR was 1.30 (95%CI 1.06, 1.59) for patients with formal social assistance and 2.13 (95%CI 1.68, 2.67) for those living in care homes.

Mortality increased with increasing levels of frailty (CFS and CCI). The mortality rate among those with oxygen requirement was 2.43 per 100 persons-day (95%CI 2.26, 2.61); there were very few deaths among those not requiring oxygen (n=11), and all these occurred in patients who did not develop breathlessness and died of an alternate cause (despite having covid-19 infection).

Table 3 shows the crude and adjusted IRR (IRRa) for the effect of wave on mortality on the same set of observations. The strongest confounders of the association in this cohort were dexamethasone, oxygen requirement, symptomatic at admission, CCI, and HFNO/CPAP. In all cases, removing the effect of the confounder (IRRa) retained the protective effect of second wave to different degrees. Mortality was 43% (95%CI 71%, 52%) lower in the second wave compared to the first wave when adjusting for the effect of dexamethasone; and 16% (95%CI 3%, 27%) lower when adjusting for oxygen requirement. Thus, oxygen requirement is acting as partial positive confounder whereas dexamethasone is acting as a negative confounder in this cohort.

**Table 3:** Crude (IRRc) and adjusted IRR (IRRa) for the effect of wave on mortality

| Covariate | N | Missing | IRRc | IRRa 95%CI | IRRa | IRRa 95%CI |
| --- | --- | --- | --- | --- | --- | --- |
| Dexamethasone | 2685 | 0 | 0.75 | (0.64, 0.86) | 0.57 | (0.48, 0.69) |
| Oxygen requirement | 2665 | 20 | 0.75 | (0.65, 0.87) | 0.84 | (0.73, 0.97) |
| Symptomatic at admission | 2685 | 0 | 0.75 | (0.64, 0.86) | 0.85 | (0.73, 0.98) |
| Charlson Comorbidity Index (CCI) | 2685 | 0 | 0.75 | (0.64, 0.86) | 0.70 | (0.61, 0.82) |
| HFNO/CPAP | 2658 | 27 | 0.73 | (0.63, 0.84) | 0.69 | (0.59, 0.80) |
| Ventilation | 2669 | 16 | 0.74 | (0.64, 0.86) | 0.78 | (0.67, 0.90) |
| Remdesivir | 2685 | 0 | 0.75 | (0.64, 0.86) | 0.76 | (0.65, 0.89) |
| ICU admission | 2685 | 0 | 0.75 | (0.64, 0.86) | 0.76 | (0.66, 0.88) |
| Tocilizumab | 2685 | 0 | 0.75 | (0.64, 0.86) | 0.73 | (0.63, 0.85) |
| Social history | 2661 | 24 | 0.74 | (0.64, 0.86) | 0.75 | (0.65, 0.88) |
| Clinical frailty score (CFS) | 2683 | 2 | 0.75 | (0.64, 0.86) | 0.76 | (0.65, 0.88) |
| Ethnicity | 2685 | 0 | 0.75 | (0.64, 0.86) | 0.74 | (0.64, 0.86) |
| BMI grouped (kg/m<sup>2</sup>) | 2421 | 264 | 0.88 | (0.74, 1.04) | 0.89 | (0.75, 1.05) |
| Age grouped (years) | 2685 | 0 | 0.75 | (0.64, 0.86) | 0.75 | (0.65, 0.87) |
| Sex | 2685 | 0 | 0.75 | (0.64, 0.86) | 0.75 | (0.65, 0.87) |

IRRc: crude RR. IRRa: adjusted RR. Covariates above the dotted line are those change in the magnitude of the effect was $\geq 5.5\%$
ICU: Intensive Care Unit, HFNO/CPAP: High Flow Nasal Oxygen/Continuous Positive Airway Pressure

In the multivariable analysis, the hazard of death during the second wave was 1.62 times higher (95%CI 1.26, 2.08) than during the first wave, after conditioning on age, sex, dexamethasone, oxygen requirement, symptoms at admission, and CCI. With age fitted as a flexible spline, and accounting for all the variables in this model, males had HR 1.21 (95%CI 1.04, 1.40); those presenting with symptoms at admission a HR 1.72 (95%CI 1.35, 2.20) and increasing CCI was (non-linearly) associated with increasing hazards of death. Dexamethasone reduced the hazard of death by 53% (95%CI 40%, 63%) when accounting for all the other factors in the model.

In the subgroup analyses of covid-19 patients requiring ICU, the hazard of death during the second wave was 2.00 (95%CI 1.10, 3.62) after conditioning on age, sex, dexamethasone, Remdesivir, Tocilizumab, and HFNO/CPAP. A summary of model development is presented in the supplemental materials.

## DISCUSSION

This cohort study examined differences in the risk of death of patients requiring in-hospital treatment for covid-19, during the first and second wave of the covid-19 pandemic in UK.

The number of covid-19 admissions was 2.05 times higher in second wave compared to the first wave (1,817 vs. 884); the proportion of admitted covid-19 patients who died was 33.6% (297 of 884) during the first wave and 25.3% (455 of 1,346) during the second wave. The crude mortality rate was 25% (95%CI 14%, 36%) lower for those admitted during the second wave compared to those admitted during the first wave (IRR 0.75 95%CI 0.64, 0.86).

We summarised the distribution of baseline characteristics at admission and medical interventions across waves for the entire study cohort (Table 1). During the second wave, younger admissions with moderate levels of frailty/CCI were more prevalent, compared to either older and/or frailer patients in the first wave. In addition to this, during the second wave, we observed an increase of covid-19 specific treatments as trial data emerged for the use of dexamethasone, Remdesivir and Tocilizumab.

The multivariable analysis attempted to account for all the available factors unequally distributed across waves and also associated with mortality (while avoiding multicollinearity in the model). We found a 1.62-fold increase in the hazard of death (95%CI 1.26, 2.08), after controlling for the effect of age, sex, dexamethasone, oxygen requirement (maximum FiO2>21%), symptoms at admission and CCI.

Dexamethasone therapy and oxygen requirement were strong confounders of the association of interest and removing either variable from the model would cause a change in the direction of the main effect. This is explained by an unequal distribution of two highly correlated covariates across waves. Oxygen and dexamethasone administration are correlated in that, the benefits of dexamethasone in the management of covid-19 hospitalised patients have been shown only for patients with hypoxaemia, but not among those with milder disease (without hypoxaemia) [21, 22]. However, this correlation, was only observed in the second wave in accordance with changes in the standard of care as evidence became available. In this cohort, the observed proportion of those who survived among those receiving both oxygen and dexamethasone was similar across waves: 75.5% (37 of 51) during the first wave, and 70.5% (866 of 1,228) during the second wave. But, as shown in Table 2, the distribution of both interventions was different across waves: in the first wave 76.3% (668 of 875) of those admitted required oxygen but 6.6% (58 of 884) received dexamethasone; in the second wave 73.5% (1,328 of 1,806) received oxygen and 68.3% (1,241 of 1,817) dexamethasone. Both variables were included in the model as the level of uncontrolled confounding reduced was larger than the error introduced due to collinear effects. After accounting for the effect of age, sex, dexamethasone, oxygen requirement, symptoms at admission, CCI and wave, dexamethasone reduced the hazard of death in this population of patients by 53% (95%CI 40%, 63%).

We further explored the effect of wave on mortality on the subpopulation of patients admitted to ICU, i.e., the most severe covid-19 patients. All these patients had oxygen therapy so, this variable was not a factor in the main model. Within this sub-group of patients, the hazard of death during the second wave was also larger than in the first wave (HR: 2.00, 95%CI 1.10, 3.62) after accounting for the effect of age, sex, dexamethasone, Remdesivir, Tocilizumab and HFNO/CPAP. This further supports the observation that risk of death in covid-19 hospitalised patients was higher in the second wave compared to the first wave, when differences in the standard of care and the characteristics of the patients were taken into account.

There is some evidence that VOC 202012/01 (alpha) is associated with increased risk of death [12-14]; but since S-gene target failure (SGTF) detection or genomic sequencing data were not available for this study population, attributing our observation of increased in-hospital mortality to variant VOC 202012/01 would largely depend on the acceptability of the assumption that said variant was dominant in our catchment area. This might not be an unreasonable assumption as prevalence of SGTF (associated with this new variant), was already at 5.8% at the beginning of November 2020, increasing sharply to reach 94.3% at the end of January 2021 [13].

### Strengths and limitations

This was a large analytical cohort study comparing groups of patients at different points in time. The overall goal was to investigate if different standards of care and possible changes in the natural history of the disease (attributed to changes in SARS-CoV-2 variants), had an impact on in-hospital mortality. We included all patients admitted to covid-19 wards for treatment.

All variables used in this study were extracted prospectively from electronic medical records ensuring data collected were the same across waves. The majority of the data were collected by experienced respiratory and ICU clinicians, and although some data inconsistencies were rectified early during data management, misclassification of covariates due transcription errors cannot be ruled out. Laboratory variables such as oxygenation parameters were obtained through the informatic department, but due to the limited quality of the electronic records, data were inconsistent and, in many cases, missing. We dichotomised this variable (FiO2) in an effort to reduce measurement error, but the coarse categorisation of oxygenation parameters into a dichotomous variable is likely to have introduced residual confounding.

Outcome and date of outcome were collected separately and ascertained from MCCD (available for 752 deaths, 94.1%). The number of deaths we observed during the first wave is consistent with numbers previously reported for the same catchment area and period [23]. However, it has been observed that during the first epidemic wave in the UK there was a larger mortality within care homes [2], so it is possible that we have underestimated the number of deaths in the first wave. This differential misclassification of outcome could have led to an overestimation of the effect of the second wave. In addition, temporal effects could also have explained some of the observed differences between waves, as fatality rates are known to be higher during winter months, when the second wave unfolded. Overall, there was a good level of data completeness with only BMI observing large numbers of missing values.

### Generalisability

This study was looking at an overall population of hospitalised adults with covid-19 in a large reference teaching London hospital. Findings are only generalisable to inpatient population.

## CONCLUSIONS

Analysis of covid-19 admissions recorded in St. Georges Hospital between 01 Jan 2020 and 31 March 2021, shows a second epidemic wave twice as large as the first one. Although crude rates would indicate a lower in-hospital mortality during the second wave; accounting for differences in the distribution of protective and risk factors (age, sex, dexamethasone use, oxygen requirement, symptoms at admission and comorbidities), suggests a higher risk of death during the second epidemic wave compared to the first. Our findings are temporally and ecologically correlated with an increased circulation of VOC 202012/01 (alpha), with estimates in agreement community-based studies. The availability of improved management and new treatments, particularly dexamethasone, was important in reducing risk of death during the second wave. This study illustrates the importance of careful clinical studies to understand risks of mortality in any future waves of covid-19.

## Data Availability

All relevant data are within the manuscript and its Supporting Information files. Data sharing and collaboration request should be directed to the corresponding author.

## CONTRIBUTIONS

We acknowledge the physicians who contributed to implementation of the study and data extraction: Dagan Lonsdale, Ashwini Maudhoo, Ashwin Sundaram, Joseph Salem, Victoria Taylor, Emma Lombard, Hannah Gardiner, Natasha Benons, Anne Dunleavy, Adrian Draper, Lois Hawkins.

