## supplemental material for "Increased risk of death in covid-19 hospital admissions during the second wave as compared to the first epidemic wave. A prospective dynamic cohort study in South London, UK"

Supplemental Table 1: Mortality rates, and crude Incidence Rate Ratios (IRRc) for all variables of interest.

| Variable / Categories | Total N=2,685 (100.0%) | Dead N=752 (28.0%) | Person-days ϕ | Rate¶ | IRRc | IRRc 95%CI | p-value |
|---|---|---|---|---|---|---|---|
| Wave | | | | | | | |
| First | 884 (100.0%) | 297 (33.6%) | 133.450 | 2.23 | Intercept | | |
| Second | 1,801 (100.0%) | 455 (25.3%) | 274.324 | 1.66 | 0.75 | (0.64, 0.86) | < 0.001 |
| Sex at birth | | | | | | | |
| Female | 1,139 (100.0%) | 274 (24.1%) | 165.514 | 1.66 | Intercept | | |
| Male | 1,546 (100.0%) | 478 (30.9%) | 242.260 | 1.97 | 1.19 | (1.03, 1.38) | < 0.001 |
| Age grouped (years)[1] | | | | | | | |
| [18,40) | 217 (100.0%) | 21 (9.7%) | 21.905 | 0.96 | Intercept | | |
| [40,60) | 678 (100.0%) | 91 (13.4%) | 92.474 | 0.98 | 1.03 | (0.65, 1.69) | |
| [60,80) | 1,067 (100.0%) | 326 (30.6%) | 176.710 | 1.84 | 1.92 | (1.27, 3.09) | |
| ≥80 | 723 (100.0%) | 314 (43.4%) | 116.685 | 2.69 | 2.81 | (1.85, 4.50) | < 0.001 |
| Ethnicity | | | | | | | |
| White | 928 (100.0%) | 270 (29.1%) | 155.554 | 1.74 | Intercept | | |
| Asian | 491 (100.0%) | 157 (32.0%) | 66.030 | 2.38 | 1.37 | (1.12, 1.67) | |
| Black | 392 (100.0%) | 114 (29.1%) | 60.200 | 1.89 | 1.09 | (0.87, 1.35) | |
| Other | 634 (100.0%) | 144 (22.7%) | 89.640 | 1.61 | 0.93 | (0.75, 1.13) | |
| Unknown | 240 (100.0%) | 67 (27.9%) | 36.350 | 1.84 | 1.06 | (0.81, 1.38) | 0.009 |
| CFS | | | | | | | |
| 1: Very fit | 195 (100.0%) | 23 (11.8%) | 21.870 | 1.05 | Intercept | | |
| 2: Fit | 290 (100.0%) | 63 (21.7%) | 41.995 | 1.50 | 1.43 | (0.90, 2.35) | |
| 3: Managing well | 898 (100.0%) | 165 (18.4%) | 124.380 | 1.33 | 1.26 | (0.83, 2.00) | |
| 4: Vulnerable | 323 (100.0%) | 94 (29.1%) | 56.970 | 1.65 | 1.57 | (1.01, 2.53) | |
| 5: Mildly frail | 263 (100.0%) | 90 (34.2%) | 42.715 | 2.11 | 2.00 | (1.29, 3.24) | |
| 6: Moderately frail | 354 (100.0%) | 138 (39.0%) | 64.909 | 2.13 | 2.02 | (1.33, 3.22) | |
| 7-9: Severely frail | 360 (100.0%) | 177 (49.2%) | 54.925 | 3.22 | 3.06 | (2.03, 4.86) | < 0.001 |
| Missing | 2 | 2 | | | | | |
| CCI | | | | | | | |
| 0 | 295 (100.0%) | 16 (5.4%) | 28.764 | 0.56 | Intercept | | |
| 1 | 288 (100.0%) | 35 (12.2%) | 35.740 | 0.98 | 1.76 | (0.99, 3.27) | |
| 2 | 334 (100.0%) | 71 (21.3%) | 51.475 | 1.38 | 2.48 | (1.48, 4.42) | |
| 3 | 347 (100.0%) | 79 (22.8%) | 57.320 | 1.38 | 2.48 | (1.49, 4.40) | |
| 4 | 369 (100.0%) | 103 (27.9%) | 59.500 | 1.73 | 3.11 | (1.89, 5.47) | |
| 5 | 348 (100.0%) | 112 (32.2%) | 58.000 | 1.93 | 3.47 | (2.12, 6.09) | |
| 6 | 297 (100.0%) | 139 (46.8%) | 49.680 | 2.80 | 5.03 | (3.10, 8.78) | |
| 7 | 188 (100.0%) | 88 (46.8%) | 30.255 | 2.91 | 5.23 | (3.16, 9.24) | |
| 8_over | 219 (100.0%) | 109 (49.8%) | 37.040 | 2.94 | 5.29 | (3.23, 9.29) | < 0.001 |
| BMI[1] (kg/$m^2$) | | | | | | | |
| <18.5 | 128 (100.0%) | 44 (34.4%) | 23.980 | 1.83 | Intercept | | |
| [18.5,25) | 784 (100.0%) | 225 (28.7%) | 134.840 | 1.67 | 0.91 | (0.67, 1.27) | |
| [25,30) | 784 (100.0%) | 197 (25.1%) | 128.134 | 1.54 | 0.84 | (0.61, 1.18) | |
| ≥30 | 725 (100.0%) | 152 (21.0%) | 107.140 | 1.42 | 0.77 | (0.56, 1.09) | < 0.001 |

| | | | | | | | |
|---|---|---|---|---|---|---|---|
| Missing | 264 | 134 | | | | | |
| Social history | | | | | | | |
| Independent | 1,566 (100.0%) | 315 (20.1%) | 217.410 | 1.45 | Intercept | | |
| Family Assistance | 503 (100.0%) | 209 (41.6%) | 85.160 | 2.45 | 1.69 | (1.42, 2.02) | |
| Formal Assistance | 357 (100.0%) | 131 (36.7%) | 69.459 | 1.89 | 1.30 | (1.06, 1.59) | |
| Care Home | 235 (100.0%) | 94 (40.0%) | 30.435 | 3.09 | 2.13 | (1.68, 2.67) | < 0.001 |
| Missing | 24 | 3 | | | | | |
| Symptomatic[2] | | | | | | | |
| No | 408 (100.0%) | 85 (20.8%) | 92.110 | 0.92 | Intercept | | |
| Yes | 2,277 (100.0%) | 667 (29.3%) | 315.664 | 2.11 | 2.29 | (1.84, 2.89) | < 0.001 |
| ICU admission | | | | | | | |
| No | 2,073 (100.0%) | 451 (21.8%) | 280.554 | 1.61 | Intercept | | |
| Yes | 612 (100.0%) | 301 (49.2%) | 127.220 | 2.37 | 1.47 | (1.27, 1.70) | |
| Oxygen requirement | | | | | | | |
| No | 683 (100.0%) | 11 (1.6%) | 104.434 | 0.11 | Intercept | | |
| Yes | 1,982 (100.0%) | 733 (37.0%) | 302.070 | 2.43 | 23.04 | (13.38, 44.53) | < 0.001 |
| Missing | 20 | 8 | | | | | |
| HFNO/CPAP | | | | | | | |
| No | 2,185 (100.0%) | 520 (23.8%) | 308.964 | 1.68 | Intercept | | |
| Yes | 473 (100.0%) | 222 (46.9%) | 92.890 | 2.39 | 1.42 | (1.21, 1.66) | < 0.001 |
| Missing | 27 | 10 | | | | | |
| Invasive ventilation | | | | | | | |
| No | 2,264 (100.0%) | 500 (22.1%) | 307.749 | 1.62 | Intercept | | |
| Yes | 405 (100.0%) | 248 (61.2%) | 97.145 | 2.55 | 1.57 | (1.35, 1.83) | < 0.001 |
| Missing | 16 | 4 | | | | | |
| Dexamethasone | | | | | | | |
| No | 1,399 (100.0%) | 376 (26.9%) | 215.994 | 1.74 | Intercept | | |
| Yes | 1,286 (100.0%) | 376 (29.2%) | 191.780 | 1.96 | 1.13 | (0.98, 1.30) | 0.173 |
| Tocilizumab | | | | | | | |
| No | 2,438 (100.0%) | 665 (27.3%) | 362.224 | 1.84 | Intercept | | |
| Yes | 247 (100.0%) | 87 (35.2%) | 45.550 | 1.91 | 1.04 | (0.83, 1.29) | 0.008 |
| Remdesivir | | | | | | | |
| No | 2,116 (100.0%) | 609 (28.8%) | 317.149 | 1.92 | Intercept | | |
| Yes | 569 (100.0%) | 143 (25.1%) | 90.625 | 1.58 | 0.82 | (0.68, 0.98) | 0.085 |

(ɸ) person-days per 100 (ǂ) mortality rate per 100 person-days

(1) Brackets indicate that the side of the interval is closed; parenthesis, that the number is excluded. (2) Symptomatic at admission

CFS: Clinical Frailty Score, CCI: Charlson Comorbidity Index, ICU: Intensive Care Unit, HFNO/CPAP: High Flow Nasal Oxygen/Continuous Positive Airway Pressure

Supplemental Table 2: Crude and adjusted IRR for the effect of wave on mortality among ICU admissions

| Covariate | N | Missing | IRRc | IRRa 95%CI | IRRa | IRRa 95%CI |
|---|---|---|---|---|---|---|
| Dexamethasone | 612 | 0 | 0.98 | (0.77, 1.24) | 2.19 | (1.30, 3.68) |
| Remdesivir | 612 | 0 | 0.98 | (0.77, 1.24) | 1.15 | (0.88, 1.50) |
| Age grouped (years) | 612 | 0 | 0.98 | (0.77, 1.24) | 0.82 | (0.65, 1.05) |
| Charlson Comorbidity Index (CCI) | 612 | 0 | 0.98 | (0.77, 1.24) | 0.84 | (0.66, 1.08) |
| Tocilizumab | 612 | 0 | 0.98 | (0.77, 1.24) | 1.07 | (0.84, 1.38) |
| Clinical frailty score (CFS) | 612 | 0 | 0.98 | (0.77, 1.24) | 0.89 | (0.70, 1.14) |
| HFNO/CPAP | 604 | 8 | 0.96 | (0.76, 1.21) | 1.03 | (0.80, 1.34) |
| Invasive ventilation | 612 | 0 | 0.98 | (0.77, 1.24) | 1.03 | (0.81, 1.31) |
| Social history | 604 | 8 | 0.97 | (0.77, 1.23) | 0.95 | (0.75, 1.20) |
| Ethnicity | 612 | 0 | 0.98 | (0.77, 1.24) | 0.97 | (0.76, 1.25) |
| BMI grouped (kg/m$^2$) | 563 | 49 | 1.14 | (0.88, 1.50) | 1.14 | (0.88, 1.49) |
| Symptomatic at admission | 612 | 0 | 0.98 | (0.77, 1.24) | 0.98 | (0.78, 1.24) |
| Sex | 612 | 0 | 0.98 | (0.77, 1.24) | 0.98 | (0.78, 1.24) |

IRRc: crude RR. IRRa: adjusted RR. Covariates above the dotted line are those change in the magnitude of the effect was ≥ 5.5%
ICU: Intensive Care Unit, HFNO/CPAP: High Flow Nasal Oxygen/Continuous Positive Airway Pressure

Supplemental Table 3: Multivariable causal models for the effect of wave on in-hospital mortality

| # | MODEL | N | Failures | Df | RMSE | SE(β) | HR | HR 95%CI |
|---|---|---|---|---|---|---|---|---|
| m1 | wave | 2,685 | 752 | 1 | -- | 0.075 | 0.75 | (0.65, 0.87) |
| m2 | wave + (A1, A2) | 2,685 | 752 | 5 | -- | 0.075 | 0.76 | (0.65, 0.88) |
| m3 | wave + (A1, A2) + (V1 - V6) | 2,646 | 738 | 18 | 0.131 | 0.131 | 1.65 | (1.28, 2.24) |
| m4 | wave + (A1, A2) + (V1 - V4) | 2,665 | 744 | 16 | 0.129 | 0.128 | 1.62 | (1.26, 2.08) |
| i1 | wave | 612 | 301 | 1 | -- | 0.120 | 0.95 | (0.75, 1.20) |
| i2 | wave + (A1, A2) | 612 | 301 | 5 | -- | 0.124 | 0.80 | (0.62, 1.01) |
| i3 | wave + (A1, A2) + (C1 -C6) | 604 | 298 | 23 | 0.323 | 0.323 | 1.93 | (1.02, 3.63) |
| i4 | wave + (A1, A2) + (C1 -C4) | 604 | 298 | 9 | 0.306 | 0.304 | 2.00 | (1.10, 3.62) |

m: model in overall population, i: model in ICU admission only (sub-analysis)
#1: crude model, #2: adjusted for a priori confounders (forced variables), #3: full model, #4: reduced model
(A1) Age fitted as flexible spline, (A2) Gender.
(V1): Dexamethasone, (V2): Oxygen requirement, (V3): Symptomatic at admission, (V4): CCI. (V5): ICU admission, (V6): HFNO/CPAP.
(C1): Dexamethasone, (C2): Remdesivir, (C3): Tocilizumab, (C4): HFNO/CPAP, (C5): CFS, (C6): CCI.
Df: model degrees of freedom, RMSE: Root Mean Square Error, SE(β) SE: of the coefficient (log scale), HR: Hazard Ratio.
